# Equivalent Binding Of Sera From Omicron And Delta Period To Future Omicron Subvariants

**DOI:** 10.1101/2023.08.04.23293670

**Authors:** Deepayan Biswas, Gokulnath Mahalingam, Rajesh Kumar Subaschandrabose, Sangeetha Priya, Rohini Ramachandran, Sevanthy Suresh, Tamil Venthan Mathivanan, Nelson Vijaykumar Balu, Kavitha Selvaraj, Arun Jose Nellickal, Pamela Christudoss, Prasanna Samuel, Ramya Devi KT, Srujan Marepally, Mahesh Moorthy

## Abstract

Throughout the COVID-19 pandemic, virus evolution and large-scale vaccination programs have caused multiple exposures to SARS CoV-2 spike protein, resulting in complex antibody profiles. Binding of sera to spike protein of future variants in the context of heterogeneous exposure, has not been studied. We tested archival sera (delta and omicron period) stratified by anti-spike levels for reactivity to omicron subvariant (BA.1, BA.2, BA.2.12.1, BA.2.75, BA.4/5 and BF.7) spike. Antibody reactivity to wild-type (CLIA) and subvariants (ELISA) spike were similar between periods (p>0.05). Both High S group and Low S group of delta and omicron periods were closely related to future subvariants by Antigenic Cartography. Anti-spike antibodies to wild type (S1/S2 and Trimeric S) clustered with subvariant-binding antibodies. Low S group interspersed between High S group on hierarchical clustering. Hybrid immunity caused by cumulative virus exposure in delta or omicron periods caused equivalent binding to future variants. Low S antibody group demonstrating similar binding is a prominent finding.

## Introduction

Continuous genetic and antigenic variability of severe acute respiratory syndrome coronavirus 2 (SARS CoV-2) has led to emergence of many variants of concern (VOCs) - Alpha (B.1.1.7), Beta (B.1.351), Gamma (P.1), Delta (B.1.617.2), and Omicron (B.1.1.529).^1,2^ Omicron, diverged initially into BA.1, BA.2, BA.3 and BA.4, BA.5 lineages, but subsequently diversified into sub-lineages (BA.2.12.1, BA.2.75, and BA.2.75.2) and recently into BA.4.6, BQ.1, BQ.1.1, BF.7 and XBB.1 causing global surges in pandemic activity.^3-5^ Large-scale vaccination programs based on the Wu Hu-1 virus spike backbone (mRNA, virus-vectored, subunit) with co-circulating virus has resulted in individuals being exposed to multiple spike proteins over the entire duration of the pandemic.

Neutralizing antibodies, a correlate of protection against SARS CoV-2 infection, prevent binding of the spike receptor binding domain (RBD) to its angiotensin converting enzyme-2 (ACE2) receptor. During infection, vaccination or reinfection, antibodies are directed towards all regions of the spike protein.^6,7^ A “hybrid” response on repeat infection/vaccination is due to activation of immune memory targeting conserved regions between the strain and vaccine.^8^ A higher level of anti-spike antibodies results in improved protection against reinfection compared to infection or vaccination alone.^8,9^ While Wu Hu-1 spike based vaccines provide repeat exposure to wild type spike, breakthrough infection with divergent strains (e.g. Omicron) stimulate rapid responses to conserved regions. Recently recommended bivalent booster vaccinations similarly stimulate recall immunity and sustain efficacy against new VOCs.^10,11^

Vaccine breakthrough causes increased intensity (level/titres) and breadth (heterologous binding) of humoral immunity to spike of antigenically distinct, ancestral strains.^12^ Infection in the pre-omicron period or omicron breakthrough (BA.1) infection provides protection against future omicron (BA.2, BA.5) infections and symptomatic disease.^13,14^ However, literature is conflicting as BA.1 breakthrough in BNT162b2 recipients provided protection against ancestral or homologous but not “future” sub-lineages BA.4 and BA.5.^15^ Taken together, infection with omicron and/or delta shows an indication of binding to “future” variant, but the mechanism is unclear.

Continued genomic surveillance over the pandemic in India and globally has led to an unprecedented amount of genetic data. However, the inherent immune-evasive nature of variants and evidence of convergent evolution among lineages, has highlighted the need for characterization of antigenic relationship between “ancestral” and newer strains.^16^ The heterogeneous and dynamic individual exposure history by infection (primary or breakthrough) and/or vaccination and the resultant diverse reactivity patterns to the spike protein determine susceptibility or protection from infection. The impact of this cumulative exposure on binding to trimeric spike protein of SARS CoV-2 omicron subvariants has not been studied to-date. Here, we probed 126 serum samples collected between April 2021 to December 2022, spanning the delta and omicron waves in India, for reactivity to wild type and omicron subvariant antigens and compared reactivity in groups stratified by anti-S (S1/S2) antibody levels. We found similarities in reactivity levels of sera, antigenic distance and clustering of sera between the delta and omicron period.

## Materials and methods

### Study design

De-identified residual sera (N=126) received at the Department of Clinical Virology for routine diagnostic testing between April 2021 and December 2022 were included in this study. Samples with collection date between April 2021 to December 2021 (N=33) and January 2022 to December 2022 (N=93) were classified as ‘Delta period’ and ‘Omicron period’ sera, respectively, representing the date range of delta and omicron VoC circulation in Vellore, as per our previous study.^12^ The study was approved by the Institutional Review Board of Christian Medical College, Vellore (IRB No. 12917).

### Detection of SARS CoV-2 antibodies

Sera were tested for antibodies to nucleoprotein (anti-N) using Elecsys anti-SARS-CoV-2 assay and to spike receptor binding domain [anti-S (RBD)] using Elecsys anti-SARS-CoV-2 S assay on a COBAS e401 analyser (Roche Diagnostics, Switzerland) based on the principle of electrochemiluminescence immunoassay (ECLIA). A cut-off index (COI) ≥1.0 was considered reactive for anti-N and ≥0.8 U/ml (units/ml) considered positive for anti-S (RBD). Antibodies to spike protein were tested using monomeric spike assay (Liaison SARS-CoV-2 S1/S2) referred to as “anti-S (S1/S2)” and trimeric spike assay (Liaison SARS-CoV-2 Trimeric S) referred to as “anti-S (TrimericS)” on the Liaison XL system (Diasorin, Italy). All samples were tested as per manufacturers’ instructions. Values ≥15 AU/ml (arbitrary unit/ml) and ≥33.8 BAU/ml (binding antibody units/ml) were considered positive for anti-S (S1/S2) and anti-S (Trimeric S) antibodies. Raw machine readouts were exported into excel spreadsheets for further analysis. Values above the upper range of detection [>400, >250 and >2080, respectively for anti-S (S1/S2), anti-S (RBD) and anti-S (TrimericS), respectively] were rounded off for the purpose of calculation.

### Detection of binding of sera to omicron subvariants by ELISA

Sera were also tested for antibodies to trimeric spike protein of omicron subvariants - BA.1, BA.2, BA.2.12.2, BA.2.75, BA.4/5, and BA.7 [purified trimeric spike protein (ACRO Biosystems) expressed in HEK-293 cells] using an in-house enzyme linked immunosorbent assay (ELISA).^12^ Briefly, 0.1 μg/well of trimeric spike protein of each subvariants were coated in high binding 96-well strip plate (Biomat, MT01F4-HB8) with incubation overnight at +4°C, followed by blocking with 3% BSA (bovine serum albumin). Sera (diluted 1/500 with 1% BSA in DPBS-T) were added to each well and incubated 1 hour at 37°C. After washing, goat anti-human IgG specific−HRP conjugate was added and incubated, followed by post-wash substrate addition (TMB). Optical density (OD) was read at 450 nm and 630 nm after addition of stop solution. Each plate included appropriate blank, negative control, positive control and internal quality control samples in addition to test sera. For each antigen, cut-off values for a sample to be considered ‘positive’ or ‘reactive’ indicative of presence of antibody (IgG) binding to trimeric spike of omicron subvariants was computed by calculating 3 standard deviations from the mean OD of negative samples (negative for anti-spike antibodies). The reactivity ratio (RR) i.e., ratio of sample OD to cut-off OD was computed for each antigen and compared between groups. Samples with RR ≥1 considered as ‘positive’ or ‘reactive’. ELISA OD and RR values were exported to excel spreadsheets for further analysis.

### Antigen cartography

Serum ELISA OD values to each of the 6 omicron subvariant antigens in the groups with collection dates between April 2021-March 2022 (Delta High, Delta Low, Omicron High and Omicron Low; N=53) were plotted in 2-D antigenic space using the Antigenic Cartography web portal (https://acmacs-web.antigenic-cartography.org/) using default settings for the data analysis. From the session output, distance matrices of sera to the 6 antigens were exported and compared across the different groups. The analysis output coordinates (antigenic map) was plotted using GraphPad Prism version 8 (GraphPad Software, USA).

### Hierarchical clustering

Further, we determined the level of hierarchical clustering of ELISA and CLIA (chemiluminescence immunoassay) data of all sera (both delta and omicron periods, N=126) against six omicron subvariants, anti-N, anti-S (RBD), anti-S (S1/S2) and anti-S (TrimericS). Data exploration and clusterability of the data was assessed with respect to assay type/antigen used and individual serum reactivity profile across all antigens (wild type and omicron subvariants). Analysis was performed using packages *(hclust, pheatmap, ggplot2)* on RStudio (v4.3.0).

### SARS CoV-2 sequences (India) and structural mapping

High quality whole genome sequences from Indian strains encompassing wild, delta and omicron infection timeline between January 2020 and December 2022 were downloaded from public repositories [N=6147 sequences (2020: 1683 sequences; 2021: 3192 sequences; 2022: 1272 sequences)]. Sequences with gaps and ambiguities were removed and unique full length spike genes extracted (N=702). Sequences were classified into wild-type and alpha (N=374), delta (N=175) and omicron (N=153) using a maximum likelihood tree generated using FastTree (v2.1.11) in Geneious Prime 2023.1.2 (Biomatters Inc.). Each of the wild, delta and omicron datasets were aligned using MAFFT (v1.0). From the translated alignments, individual subdomains – S1 N terminal domain (NTD) (AA 16-303), S1 receptor binding domain (RBD) (AA 319-541) and S2 subunit (AA 686-1273) were extracted using the NC_045512 (SARS CoV-2 isolate Wuhan Hu-1; AA 1-1273) and extent of variability was compared within and between the VOCs. The amino acid conservation across the 3 variants were plotted on the Wu Hu-1 spike protein structural backbone (PDB: 6XRA) to illustrate conservancy across all variants in the S1 and S2 domains.

### Statistical analysis

Levels of anti-spike IgG across variants was analysed by nonparametric one-way ANOVA (Friedman test). Statistical analysis and visualization of data was performed in GraphPad Prism version 8 (GraphPad Software, USA). Mann Whitney test was used to analyse significance between groups.

## Results

This study was conducted on samples collected between April 2021 and December 2022 (21 months) covering the delta and omicron waves at Vellore in South India (Figure 1). Median (IQR) levels of anti-N, anti-S (RBD), anti-S (TrimericS) and anti-S (S1/S2) were 33 (112), 251 (0), 712 (1914) and 270 (313) respectively. Of the 126 samples collected, 33 (26.2%) were from delta period while 93 (73.8%) were from the omicron period. Among the 126 sera tested, positivity for anti-N, anti-S (RBD), anti-S (TrimericS) and anti-S (S1/S2) was 58% (19/33), 100% (33/33), 97% (32/33) and 100% (33/33) in the delta period and 87% (81/93), 100% (93/93), 98% (91/93) and 100% (93/93) in the omicron period, respectively. Median (IQR) antibody levels in the delta period were significantly lower (Mann Whitney test, p<0.05) compared to the omicron period for anti-N [8.6 (81.6) versus 42.3 (118.6)], anti-S (TrimericS) [270 (767) versus 981 (1864)] and anti-S (RBD) [251 (72.7) versus 251 (0)]. Of the 126 sera tested, only 107 were positive (RR≥1) to at least 1 of 6 trimeric omicron subvariants spike antigens. Of these 107, 93 were reactive to all 6 omicron antigens (P_6_), while 14 were reactive to any 1 antigen (P_any_). The 19 samples negative (RR<1) to all antigens were excluded from analysis.

**Figure 1.**
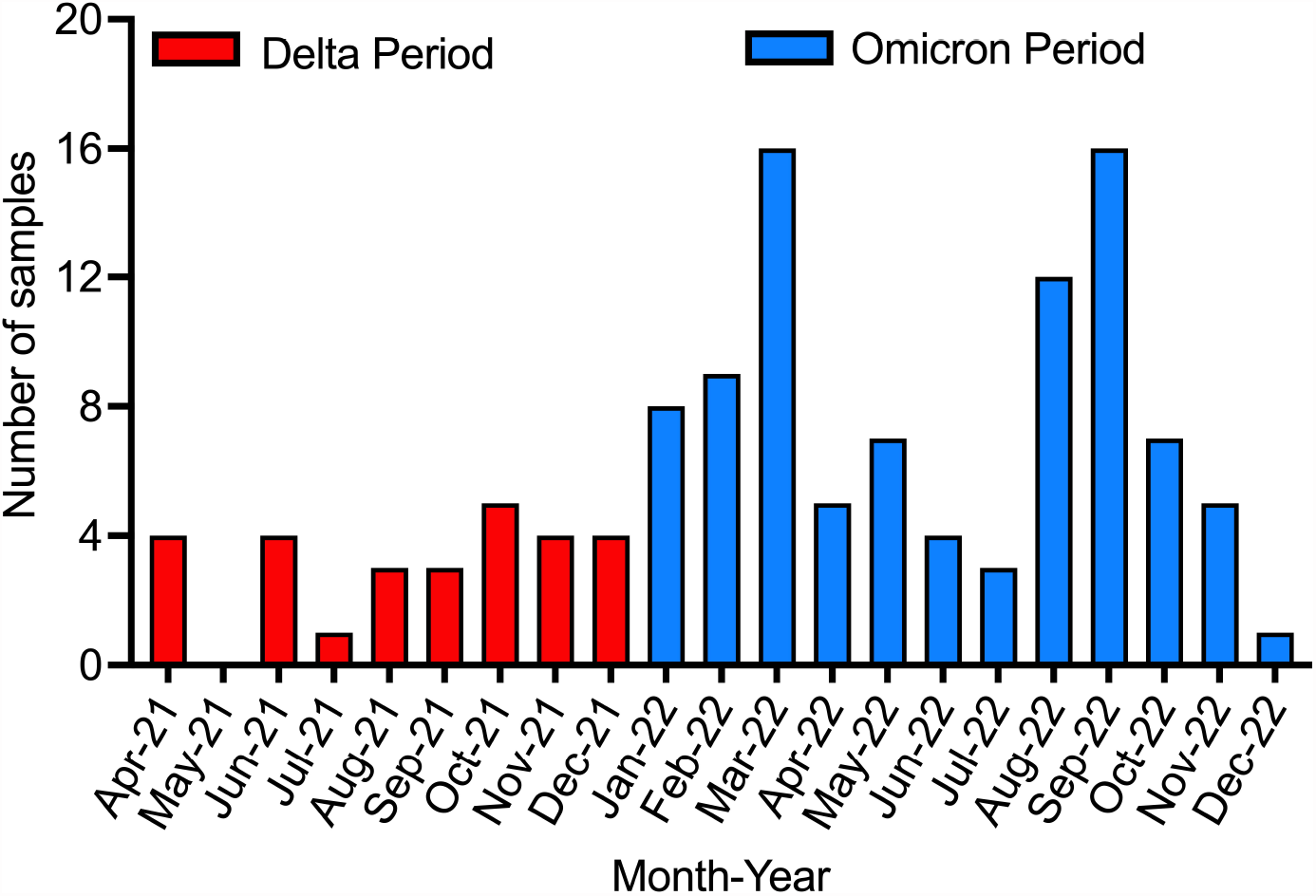
Serum samples collected between April 2021 to December 2021 (Delta period) and January 2022 to December 2022 (Omicron period). X-axis represents month and year. Y-axis represents number of serum samples per month. Delta (B.1.617.2) and Omicron waves were based on sequencing data. Blood collected 4-6 weeks after SARS CoV-2 PCR confirmation from respiratory sample.

Sera were stratified in each period into “High S” or “Low S”, based on levels above or below the mean anti-S (S1/S2) antibody (245 AU/ml) levels, respectively. We compared antibody levels in the “Delta High” (N=9) with “Omicron High” (N=58) and “Delta Low” (N=16) with “Omicron Low” (N=24) groups. The percentage of P_6_ reactivity was 100% (9/9) and 95% (55/58) among Delta High and Omicron High, respectively while it was 62% (10/16) and 79% (19/24) for Delta Low and Omicron Low, respectively.

The levels of anti-N, anti-S (RBD) and anti-S (Trimeric S) and RR to omicron antigens (BA.1, BA.2, BA.2.12.1, BA.2.75, BA.4/5 and BF.7) is shown in Figure 2. Median (IQR) levels of anti-N, anti-S (RBD) and anti-S (TrimericS) antibodies (Fig 2A-C) were similar within the Low S (‘Delta Low’ and ‘Omicron Low’) and High S (‘Delta High’ and ‘Omicron High’) groups, with the exception of anti-N in the Low S [0.6(64.5) and 35.2(92.9) in Delta Low and Omicron Low, respectively, p=0.031]. Next, we compared reactivity/binding levels to six trimeric spike proteins of omicron subvariants (BA.1, BA.2, BA.2.12.1, BA.2.75, BA.4/5 and BF.7) between the High S - “Delta High” (N=9) and “Omicron High” (N=58) as well as the Low S - “Delta Low” (N=16) and “Omicron Low” (N=24) groups. Using BA.1 as reference, we noted that for both Delta High and Omicron High groups (Figure 2D and 2E), we noted a lower antibody level to other omicron variants (Friedman test, p<0.05) with the exception of BA.2.75 (p>0.05) in the Delta High group. However, in the Low S group (Figure 2G and 2H), similar reactivity as BA.1 (p>0.05) was seen to BA.4/5 variant (among Delta Low) and BA.2, BA.2.12.1 and BA.2.75 variants (among Omicron Low). We then compared the reactivity to each antigen across the High S and Low S groups. The median range of reactivity of Delta High was 2.3-3.8 and that of Omicron High was 2.3-3.5. The median (IQR) RR value to each of the 6 antigens were similar between Delta High and Omicron High indicating similar reactivity to the antigens [p>0.05, Mann Whitney test; Figure 2F]. Further, we examined the Low S sera and found a median range of 1.3-1.6 and 1.5-1.7 for Delta Low and Omicron Low respectively. Likewise, the High S groups, levels were similar for all antigens between the Low S groups [p>0.05, Mann Whitney test; Figure 2I].

**Figure 2.**
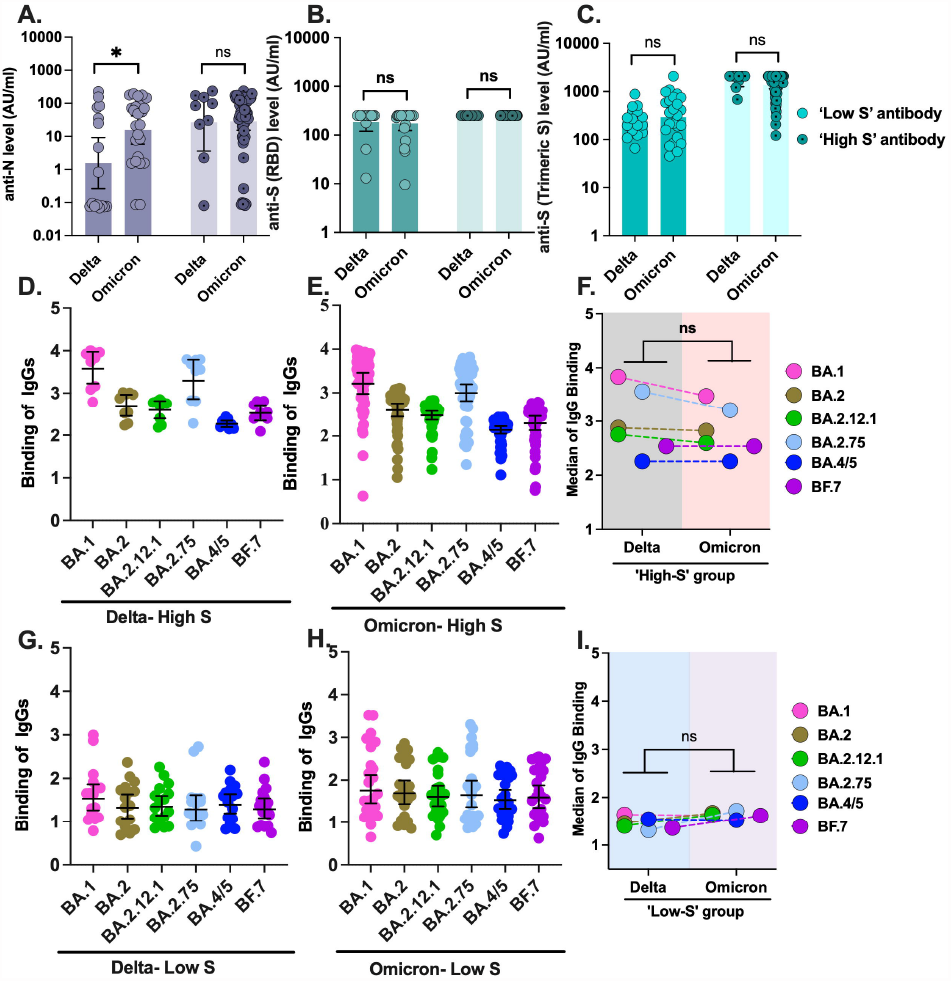
SARS CoV-2 antibody levels to spike of Wild-type and Omicron subvariants. 2(A-C) SARS CoV-2 anti-nucleoprotein and anti-spike (RBD and TrimericS) antibody levels across delta and omicron variants (both ‘High group’ and ‘Low group’). X-axis shows Delta Low, Omicron Low, Delta High and Omicron High sera. Y-axis shows anti-nucleoprotein (2A), anti-spike (RBD) (2B) and anti-spike (TrimericS) (2C) antibody levels in AU/ml. 2(D-I) Reactivity/binding levels to 6 trimeric spike proteins of omicron subvariants (BA.1, BA.2, BA.2.12.1, BA.2.75, BA.4/5 and BF.7) in the High S - “Delta High” (N=9) and “Omicron High” (N=58) (2D and 2E) as well as the Low S - “Delta Low” (N=16) and “Omicron Low” (N=24) groups (2G and 2H). Comparison of levels of anti-S (TrimericS) antibody across each omicron subvariants between Delta High and Omicron High (2F) and between Delta Low and Omicron Low (2I). X-Axis shows reactivity of 6 different omicron variants by delta sera and omicron sera. Y-axis shows levels of anti-S antibodies in AU /ml (2D, 2E, 2G and 2H) and median IgG binding (2F and 2I). The omicron variants are coloured as pink (BA.1), brown (BA.2), green (BA.2.12.1), sky blue (BA.2.75), blue (BA.4/5) and purple (BF.7). Level of statistical significance is denoted by * for p <0.05, ** for p < 0.01, *** for p < 0.001, not significant (p>0.05) if not indicated.

### Delta and omicron sera show similarity in antigenic space

Given the heterogeneity in reactivity (P_6_ and P_any_) and similar levels seen between High S (Delta High and Omicron High, Figure 2F) and Low S (Delta Low and Omicron Low, Figure 2I) groups, we quantified antigenic distances between sera using antigenic cartography. We included 53 serum samples between April 2021-March 2022. Of these, 25 were from the delta period (9 ‘Delta High’ and 16 ‘Delta Low’) and 28 from omicron period (21 ‘Omicron High’ and 7 ‘Omicron Low’). The antigenic maps for ‘High S’ and ‘Low S’ groups as well as mean (SD) distance of sera to the 6 omicron variant antigens are shown in Figure 3A-D. It is noteworthy that all of sera, were placed within a grid of 1 antigenic unit with respect to all antigens. This was seen for both High S (Figure 3A) and Low S (Figure 3B) groups. Further, the distance of individual sera in the Delta High and Omicron High (Figure 3C) to each of the antigens was similar between the groups (ANOVA, p>0.05). The distance of the Low S groups (Figure 3D) from the antigens was also similar (ANOVA, p>0.05). These findings indicate that delta and omicron sera were closely related to the omicron subvariants in antigenic space.

**Figure 3.**
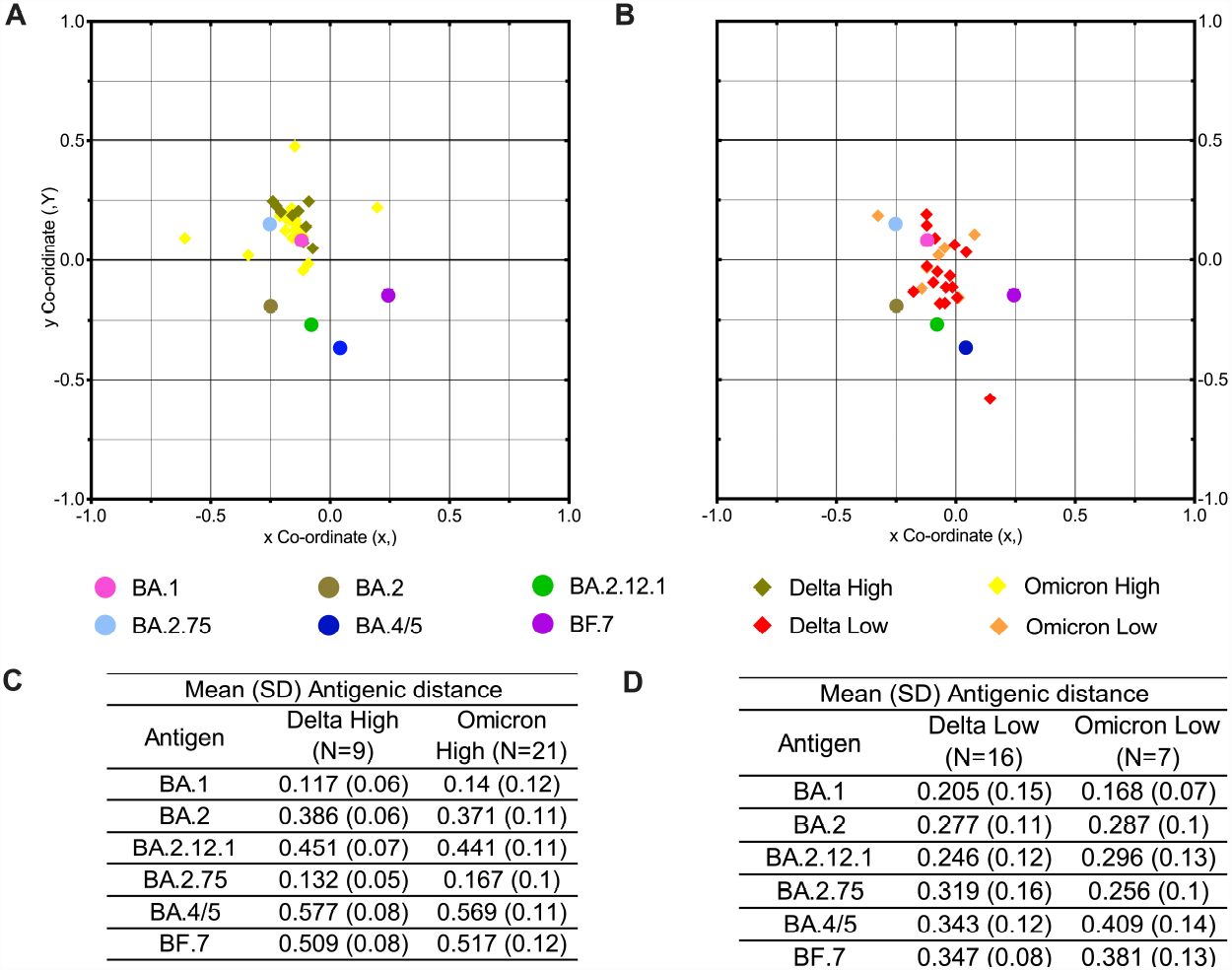
Antigenic cartography of Delta and Omicron sera against Omicron subvariants. Antigenic Cartography for the ‘High S’ (Delta High and Omicron High) (3A) and ‘Low S’ (Delta Low and Omicron Low) (3B) sera groups against six omicron subvariants in 2D antigenic space. Omicron subvariants are shown as circles and sera are shaped as diamond. Each diamond corresponds to sera of one individual. Both axes of the map are antigenic coordinates. Mean (SD) antigenic distance in between the ‘High S’ groups (3C) and ‘Low S’ groups (3D) against six omicron variants. Sera are coloured as olive (Delta High), gold (Omicron High), red (Delta Low) and orange (Omicron Low). The omicron variants are coloured as pink (BA.1), brown (BA.2), green (BA.2.12.1), sky blue (BA.2.75), blue (BA.4/5) and purple (BF.7). The distance between points in the map can be interpreted as a measure of antigenic similarity, where the points more closely together indicate that they are antigenically more similar.

### Hierarchical clustering

Given the observed antigenic distance but heterogeneity in levels/reactivity of sera groups with respect to anti-N, anti-S (RBD), anti-S (TrimericS), anti-S (S1/S2) and omicron subvariant antigens, we performed a hierarchical clustering of all sera (N=126) based on results of CLIA assays or OD values to each antigen (Figure 4). We further noted, heterogeneous spike binding manifested as positivity to 6 omicron variants (P_6_, N=93) or any one (P_any_, N=14) or none at all (negative, N=19) in delta and omicron periods. High S sera showed completely P6 while Low S sera showed a mix of P_6_, P_any_ and negative. Hierarchical clustering of antibody testing (columns) showed distinct clusters with no delineated segregation by serum group or reactivity. Sera from the Low-S groups clustered together but were also seen to be interspersed with Delta High and Omicron High. Particularly notable was the finding of sera demonstrating P_6_ reactivity to have High S (Delta High and Omicron High) and Low S (Delta Low and Omicron Low) groups interspersed. The clustering of assay type (rows), revealed clustering of omicron variants ELISA results together along anti-S (S1/S2) and anti-S (TrimericS) branches, while the anti-S (RBD) and anti-N were on separate branches.

**Figure 4.**
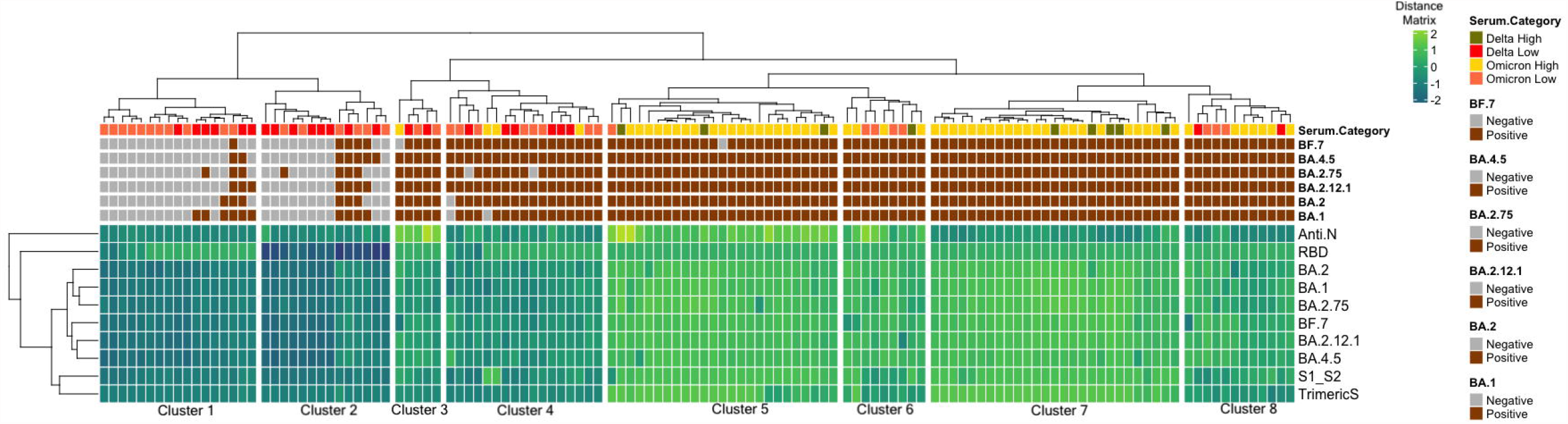
Hierarchical clustering of Delta and Omicron sera against spike of Wild-type and Omicron subvariants. Hierarchical clustering of all sera (N=126) based on results of the CLIA assays or OD values against six omicron subvariants, anti-N, anti-S (RBD), anti-S (S1/S2) and anti-S (TrimericS). The dendrogram was annotated as per High S and Low S group and positivity (categorical) to individual antigens. The clusterability (Simpsons index) was 0.73. The optimal number of clusters based on k-means distance was 8. The heatmap is split according to clusters 1-8 from left to right by white space. Clusters grouped together based on ELISA antigens (i.e. P6 or Pany). Analysis was performed using packages (*hclust, pheatmap, ggplot2*) on RStudio (v4.3.0). Sera from delta and omicron period (both ‘High S’ and ‘Low S’ groups) are coloured as olive (Delta High), gold (Omicron High), red (Delta Low) and orange (Omicron Low). Individual sera positive to any omicron subvariant are indicated with brown colour and negative to all omicron subvariant are indicated with grey colour. Distance matrix ranged from -2 to +2.

### Amino acid conservation in SARS CoV-2 sequences from India

S1 and S2 domain of the downloaded SARS CoV-2 sequences were analysed to assess level of variability. Through our analysis approach, we included high quality publicly available sequence data from India to capture the entire spectrum genetic diversity of SARS CoV-2 in India between 2020 to 2022. Overall, percentage identity at amino acid level across all variants of SARS CoV-2 was 63.5% (185/291), 80.3% (179/223) and 83.2% (489/588) in the S1-NTD, S1-RBD and S2 domain. Higher % conservation was seen in the fusion peptide (FP 1 and 2) at >90%. The % conservation ranged from 75-92%, 87-94% and 81-96% within wild, delta and omicron lineages, respectively, with higher conservancy seen among S2 compared to S1 regions. The conserved regions of S1 and S2 (trimeric configuration of PDB: 6XRA) with highlighted broadly neutralizing epitopes from literature are shown in Figure S1.

## Discussion

In this study, we examined cross-reactivity of sera from delta and omicron period (N=126) to spike proteins of wild type Wu Hu-1 and six omicron subvariants. We stratified these sera based on level of anti-S [mean anti-S (S1/S2) = 245 AU/ml] into ‘High S’ (≥245 AU/ml) and ‘Low S’ (<245 AU/ml). High S may be due to hybrid immunity i.e., exposure to multiple viruses and/or vaccines with immune memory boosting and ‘Low S’ due to primary infection or vaccination. We excluded samples (N=19) with reactivity ratio (RR)<1 to all the six omicron variants and compared antibody levels between the ‘High S’ and ‘Low S’ groups.

### Omicron and delta sera show similar reactivity to omicron subvariants and wild-type antigen

In the ‘High S’ groups (i.e. Delta High and Omicron High), reactivity ratio (RR) of sera to the omicron subvariant antigens (BA.1, BA.2, BA.2.12.1, BA.2.75, BA.4/5 and BF.7) ranged from 2.1-4.0 and 0.6-3.9 for Delta High and Omicron High, respectively, and these were found similar (Mann-Whitney U-test, p>0.05). These groups showed similar reactivity levels to ‘wild-type’ spike antigens - anti-S (RBD), anti-S (TrimericS) as per manufacturers’ assay configuration for these commercial kits (CLIA/ECLIA). This indicates antibodies detected by the above methods were similar or had common binding regions.

Delta High groups represent a cumulative immunity to wild type and delta while Omicron High groups, represent cumulative exposure to wild type, delta and omicron and both groups include breakthrough infections among the vaccinated, respectively. Delta High sera were collected prior to the emergence of omicron in India (January 2022) and evidence of binding to omicron subvariant spike points to similarity or commonality of binding region between all variants (wild type, delta and omicron). India experienced multiple omicron waves.^3,17^ Reinfections were common in the omicron period^18^ and individuals would likely have had multiple exposures to omicron subvariants (BA.1, BA.2, BA.2.12.1, BA.4/5, BA.2.75.2 etc.)^19,20^ and thus can be expected to bind to conserved spike regions across omicron sub-lineages. Similarly, in the Low S groups, Delta Low and Omicron Low demonstrated similar RR level between them (Mann Whitney U-test, p>0.05). Likewise, levels were also similar between groups to wild type antigens. Sera in Low S groups, may be considered primary infection, and the observed binding to ancestral and “future” variants suggests antibody repertoire binding to S1 and S2 domains conserved across SARS CoV-2 wild type, delta and omicron variants. Re-infection or breakthrough infection leads to increased depth (antibody levels) as well as breadth (reactivity to heterologous viruses) of spike reactivity.^12,21^ In the time course of repeat infections/exposures, S1 particularly NTD shows antigenic divergence between wild-type and successive VoCs, but S2 is relatively conserved^22,23^ and contributes to serological cross reactivity.

### Delta and omicron sera overlap in antigenic space

To examine reactivity to “future” antigens, antigenic cartography was performed in 2D space using ELISA OD values on sera collected between April to December 2021 (N=25, Delta) and January to March 2022 (N=28, Omicron). The dates of blood collection for these selected sera pre-dates the emergence of the viruses - BA.2.12.1, BA.2.75, BA.4/5 and BF.7 which emerged sequentially after May 2022. For Delta High and Omicron High groups, mean distance of sera to antigens ranged from 0.18-0.58 units and 0.14-0.57 units respectively, and were similar between groups. Similarly, in the Low S sera, both groups occupied one unit of antigenic space and mean distances to the 6 antigens were similar (Delta Low range: 0.21-0.35 units; Omicron Low range: 0.17-0.41 units). All sera (both High S and Low S groups) were placed within a narrow antigenic window of 1 unit in relation to subvariant antigens.

Wild, delta and omicron VOCs are placed wide apart due to amino acid mutations resulting in escape from neutralization.^24-26^ Antigenic distance is a measure of relatedness between sera and antigens and use multidimensional scaling to generate consensus relatedness in 2D space. As a result, sera raised from homologous viruses do not occupy exact overlapping coordinates, but exist as a cluster. Our finding of aggregation of serum samples in a narrow antigenic space, indicated a close relationship and binding to similar regions on the spike. S2 domain of spike is more conserved compared to S1.^22,27^ Taken together, these suggest that the binding regions of the sera are antigenic sites conserved between the variants.

### Antibody reactivity profiles to wild and omicron antigens across entire time period of the pandemic

We examined anti-S (wild type) and anti-S (omicron subvariant) reactivity by hierarchical clustering in a Euclidean distance framework. Our observed clustering based on antigen type employed, showed grouping of non-RBD spike antigens represented by S1/S2 and TrimericS with omicron subvariant antigens. Further, we noted a dissimilarity of anti-S (RBD) and anti-N, which are based on wild type antigens with other spike antigens. These suggest that the “future variant” binding antibody repertoire target the conserved, consensus regions across wild and omicron spike but excluding RBD.

Delta Low and Omicron Low sera were interspersed with Delta High and Omicron High among clusters 3-8 (Figure 4). It suggests similarity in reactivity profile. Low S groups, due to primary infection result in reactivity to S1 and S2 subunits of homologous viruses. The reactivity to all antigens (P_6_) seen with both Delta Low and Omicron Low sera, can be explained by antibody binding to regions on the spike common between delta and omicron. Hybrid immunity seen in the High S group, showed mainly P_6_ reactivity, and is likely due to boosted level of antibodies to conserved regions. We have previously shown that omicron breakthrough results in higher binding to predecessor or ancestral viruses, mediated by antibodies binding to conserved regions.^12^

The infection/exposure history and impact on binding to future variants seen with SARS CoV-2 is analogous to sequential vaccination with a chimeric haemagglutinin (cHA). Booster with a divergent HA head (HA1, heterologous) after a primary (homologous) vaccination induces antibodies to stem (HA2) of influenza A viruses.^28^ The subdominant stalk is retargeted by immune memory from the primary vaccine and boosted levels of stem-specific antibodies mediate protection against heterologous challenge.^29^ Similar to influenza, variability of SARS CoV-2 spike is mainly in the S1-NTD compared to S2.

Serological profiles especially the High S group, need further experimental validation of binding specificity. Further studies into these antibodies for SARS CoV-2 may help the development of assays that can measure the efficacy of pan-coronavirus vaccine candidates, which can mediate protection from future variants as well as broadly-neutralizing (bn) antibodies directed in the conserved S2 domain could that offer cross-variant and pan-genus protection. Fusion peptide (FP) and heptad repeat 2 (HR2) sites may induce production of more broadly active nAbs. These same classes of antibodies have been shown to be involved in protection from severe disease.

### Public Health impact

Our study showed that antibody binding to future omicron variants was seen to occur even among sera without exposure to these variants. These may partly explain the lack of sustained transmission in India which had a severe epidemic of delta but also implemented a successful vaccination program. Similarly, this existing immunity may prevent future epidemics. This would be of great benefit to populations in whom COVID-19 vaccine is not yet approved for use, i.e. children <12 years of age.

### Limitations of the study

Our study used de-identified sera instead of an ideal setting of a temporal follow-up cohort with detailed documentation of infection and vaccination. Confirmation of results with a S2-specific assay would further strengthen the conclusion of this study. The inferences thus represent a sum-total of reactivity to conserved regions. Antigenic cartography depicts the antigenic distance based on neutralizing antibodies. In our study, we used ELISA OD values and not neutralizing end-point titres, potentially leading to inaccurate distance estimation between the omicron variants.

## Conclusion

Complex antibody profiles generated in the background of time-varying, virus exposure and/or vaccination determine reactivity to antigens of a rapidly evolving virus. An immune profile of high anti-spike levels generated by hybrid immunity from cumulative exposure to SARS CoV-2 in delta or omicron periods, show equivalent binding to “future” variants. Furthermore, a ‘Low S’ profile due to primary infection demonstrating binding to subvariants is of prominence. These could potentially inform pan-coronavirus vaccine development, similar to HA-stem binding antibodies generated by universal influenza vaccines.

## Acknowledgements

Mahesh Moorthy acknowledges Department of Biotechnology, India for financial support through BT/PR40390/COT/142/1/2020.

Srujan Marepally acknowledges Department of Biotechnology, India for the financial support through grants, BT/PR25841/GET/119/162/2017; and BT/PR40446/COV/140/5/2021.

## Data Availability Statement

Original data will be made available upon request.

## Ethical Approval

The Institutional Review Board of Christian Medical College, Vellore (IRB No. 12917). No experimentation was done on human subjects. Waiver of consent was obtained for all the tests conducted in the study, where we used de-identified residual sera.

## Author contributions

Conceptualization: Mahesh Moorthy (*M*.*M*.), Srujan Marepally (*S*.*M*.); Data Curation: Deepayan Biswas (*D*.*B*.), Rajesh Kumar Subaschandrabose (*R*.*K*.*S*.), Gokulnath Mahalingam (*G*.*M*.), Rohini Ramachandran (*R*.*R*.), Prasanna Samuel (*P*.*S*.); Formal Analysis: *D*.*B*., *R*.*K*.*S*., *G*.*M*., *P*.*S*.; Funding Acquisition: *M*.*M*.; Investigation: *D*.*B*., *G*.*M*., *R*.*K*.*S*., *R*.*R*., Sangeetha Priya (*S*.*P*.), Tamil Venthan Mathivanan (*T*.*V*.*M*.), Arun Jose Nellickal (*A*.*J*.*N*.), Pamela Christudoss (*P*.*C*.), Sevanthy Suresh (*S*.*S*.), Ramya Devi KT (*R*.*D*.*K*.); Methodology: *D*.*B*., *G*.*M*., *R*.*K*.*S*., *S*.*P*., *R*.*R*., *T*.*V*.*M*., Nelson Balu Vijaykumar (*N*.*B*.*V*.), Kavitha Selvaraj (*K*.*S*.); Project Administration: *M*.*M, S*.*M*.; Resources: *M*.*M, S*.*M*.; Software: *P*.*S*., *M*.*M*.; Supervision: *M*.*M*., *S*.*M*.; Validation: *G*.*M*; Visualisation: *D*.*B*., *G*.*M*., *R*.*K*.*S*., *R*.*R*.; Writing original draft: *D*.*B*., *R*.*K*.*S*., *G*.*M*.; Writing review and editing: *All authors*.

