## Supplementary figures and images for "Equivalent Binding Of Sera From Omicron And Delta Period To Future Omicron Subvariants"

### Figure S1

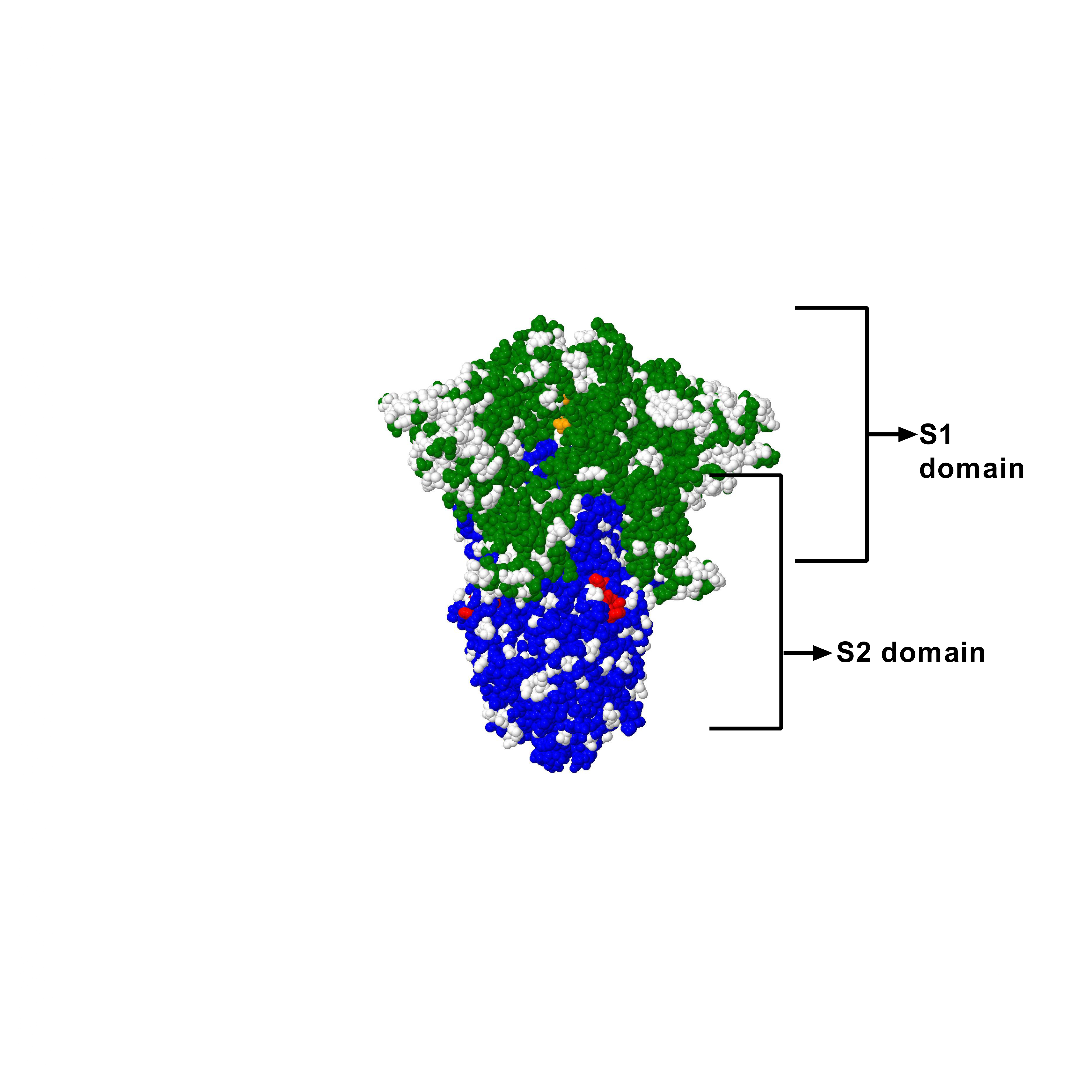
