## Supplementary material for "Equivalent Binding Of Sera From Omicron And Delta Period To Future Omicron Subvariants": Figure S1 legend and Table S1

**Title:**

**Legend for Supplementary Figure 1:**

Structure of the trimeric spike protein of Wu Hu-1 SARS CoV-2 taken from NCBI Protein ID sequence YP 009724390 (Gene ID: 43740568) (PDB number: 6XRA). The structure is derived using Swiss Model software (The Centre for Molecular Lifesciences, University of Basel) *[Waterhouse et al. SWISS-MODEL: homology modelling of protein structures and complexes. Nucleic Acids Res. 46, W296-W303 (2018).].* Here the spike protein is composed of two domains- S1 (AA position: 14-685) and S2 (AA position: 686-1144). Green colour indicates conserved regions of S1 domain. Blue colour indicates conserved regions of S2 domain. White colour in between represents the non-conserved sites of spike protein. The fusion peptide in S2 domain of spike is marked (red colour; AA position 816-823) with ‘broadly neutralizing’ antibody epitopes [Monoclonal antibodies (COV 44-62, COV 44-79 and COV 91-27) taken as reference *(Dacon C et al. Science. 2022)*]. The NTD in S1 domain of spike is marked (orange colour; AA positions 369, 377, 380, 383, 385) with ‘broadly neutralizing’ antibody epitopes [Monoclonal antibody (S2X259) taken as reference *(M Alejandra Tortorici et al. Nature 2021)*].

**Supplementary Table 1:**

Accession numbers of whole genome SARS CoV-2 sequences from India encompassing wild, delta and omicron infection timeline between January 2020 and December 2022 (refer methods) were used for structural mapping (N=702). These sequences were classified into wild-type and alpha (N=374), delta (N=175) and omicron (N=153).

Wild-type and Alpha Variants (N=374):

| MT800515 | MZ336025 | MT481898 | MW425837 | MT800818 |
| --- | --- | --- | --- | --- |
| MZ262295 | MZ261729 | MZ292135 | OM073956 | MT801051 |
| MT953981 | MT800515 | MT635856 | MZ268643 | MZ268208 |
| OM073945 | MW595950 | MT800939 | MZ318038 | MT509501 |
| MT012098 | MT759588 | MZ268636 | MZ562751 | MZ562754 |
| MT435085 | MZ254837 | MZ336033 | OM073904 | MW819940 |
| MZ262291 | MW600455 | MW165862 | MT741490 | MT594112 |
| MW181840 | MT940464 | MT951393 | MT607612 | MT481895 |
| MW425837 | MT953887 | MT576048 | MW191501 | MT941276 |
| MZ254908 | MW242671 | MZ254877 | MT467261 | OM073862 |
| MW828341 | MT741096 | MZ325260 | MT740619 | MZ342810 |
| MT435083 | MW819619 | MW243003 | MZ268208 | MW969757 |
| MZ723921 | MZ268644 | MT509650 | MZ269101 | MT415320 |
| MZ292150 | OM073924 | MZ317918 | MZ314964 | MT451881 |
| MT050493 | MZ262335 | OM915415 | MZ317920 | MZ262303 |
| MT664197 | MT953890 | MT467253 | MW242708 | MT666042 |
| MT950349 | MT664727 | MT451886 | MZ340545 | MT451880 |
| MT666042 | OM073922 | MT953980 | MT050493 | MT758683 |
| MZ227612 | MZ314980 | MZ292146 | MZ261819 | MT953893 |
| MZ269094 | MZ292128 | OM915406 | MT806176 | MZ267529 |
| MZ261728 | MT953893 | MZ269079 | MT607602 | MW191511 |
| MW242706 | MW600461 | MW645477 | MZ254876 | MT941059 |
| MT801048 | MW242999 | OM073911 | MZ336028 | MZ262303 |
| MZ254903 | OM915402 | MT800024 | OM073843 | MW595948 |
| MZ261927 | MT940462 | MT451882 | MT740441 | MZ021503 |
| MW165857 | OM073851 | MT758197 | MW600455 | OM759831 |
| MZ310509 | MZ266568 | MW173463 | MZ317906 | MZ342809 |
| MT772253 | MZ254770 | MZ269242 | MT740706 | MZ227545 |
| MT509505 | MW165857 | MT940461 | MT758439 | MT509509 |
| MT758193 | MZ254837 | MT740708 | MT954959 | MW595919 |
| MZ269079 | MT467243 | MT560706 | MT940478 | MZ268634 |
| MZ268599 | MZ318038 | OM902057 | OM073884 | MT675950 |
| OM073899 | MW191511 | MT740723 | MZ254902 | MT940472 |
| OM073932 | MT451877 | MT451881 | MT954067 | MZ292133 |
| MT759714 | MZ269095 | MT607612 | MW828341 | OM073931 |
| MT415321 | MT509497 | MZ268209 | MT509497 | MZ544380 |
| MZ227612 | MZ960573 | MZ262291 | MZ292792 | MZ317907 |
| MT509501 | OM915415 | MZ261734 | MZ342808 | MZ255007 |
| MZ292157 | MW242964 | MW166214 | MZ292154 | MT951181 |
| MT496976 | MT539166 | MT467250 | MZ292127 | MW191512 |
| MZ266543 | MW242965 | MZ292130 | MZ268600 | MZ262298 |
| MZ262334 | MT758375 | MT940452 | OM073871 | MZ254992 |
| MT451890 | MZ336023 | MT635405 | OM073877 | MZ562762 |
| MZ340537 | MT467247 | MW596415 | NC_045512 | MZ261895 |
| MT560682 | MZ314964 | MT940476 | MT940451 | OM073826 |
| OM073869 | MZ314700 | MT800995 | MZ702552 | MW165876 |
| MT451888 | MZ254839 | MZ562762 | MT467240 | MZ342591 |
| MZ254774 | MZ310508 | MW595982 | MT560689 | MT758439 |
| MT940451 | MZ268634 | MZ267528 | MT483560 | MW181804 |
| MT801048 | MW590718 | MT415322 | MT800002 | MW600654 |
| MW181848 | MW590723 | MZ310512 | MZ266553 | MW969752 |
| MT358637 | MT676388 | MZ227545 | OM073881 | MZ261635 |
| OM900175 | MZ268601 | MT953991 | MT496993 | MT800013 |
| MT740717 | MT467251 | MW181834 | MW595948 | MZ292146 |
| OM073825 | MZ292136 | OM759836 | OM073848 | MZ292134 |
| MZ292150 | MZ261812 | MT800041 | MT576033 | MT772297 |
| MZ262296 | MT941272 | MT953978 | MT496989 | MZ269147 |
| MT496987 | MT954959 | OM073826 | MT664201 | MW166221 |
| MT772271 | MZ342809 | MZ261914 | MT576061 | MT451889 |
| MW969758 | MW242964 | MT772271 | OM073825 | MZ292137 |
| MT940473 | MZ562751 | MT416725 | MW242653 | MT467252 |
| MZ254787 | MW165869 | MW173460 | OM915402 | MZ562755 |
| MZ268602 | MZ317890 | MZ292138 | MZ261726 | MW181838 |
| MZ292128 | MZ724490 | MW425563 | MT664727 | MT635855 |
| MW645476 | MT415320 | MW969753 | MZ254903 | OM073936 |
| MW927136 | MT576043 | MW181811 | MT635272 | MZ268209 |
| MZ356567 | MZ268850 | MT560704 | MT953891 | MZ254875 |
| MT800758 | MT800995 | MZ342808 | MT496979 | MW645476 |
| MT539176 | MT451876 | MZ254770 | MZ266543 | MZ723921 |
| MZ292135 | MZ292153 | MT576039 | MZ021503 | OM073919 |
| MT951393 | MZ340545 | MT954071 | MZ292138 | MZ340511 |
| MT941056 | MZ325261 | MT940472 | MT676366 | MT416726 |
| MT560706 | MT496996 | MZ292152 | MZ292094 | MT496988 |
| MZ336032 | MW828340 | MZ262293 | MT799974 | MZ317915 |
| MZ256065 | MZ340512 | MT509509 | MW181837 |  |

Delta Variant (N=175):

| OK083551 | OK189617 | ON052763 | OM899797 | OM847392 |
| --- | --- | --- | --- | --- |
| MZ713400 | OP599887 | OK356472 | OP430922 | OP430917 |
| OM899797 | OK189652 | OK356463 | OK190631 | OM883832 |
| OM899974 | OK189614 | OK356461 | OK190649 | OM884010 |
| MZ413298 | OM884047 | ON052774 | OK356412 | OM884005 |
| OL966453 | OK085466 | OM884064 | MZ558154 | OK061186 |
| OK356460 | OM899974 | OK190633 | OK356417 | OK067232 |
| MZ558161 | OK356416 | OM918219 | OK067272 | OM884010 |
| OK356466 | OK190617 | OK189636 | OK189653 | OK189634 |
| OK356427 | OK356459 | OK189625 | OK189637 | OK356454 |
| OK189611 | OK356413 | OK190638 | OM884045 | OM884049 |
| OK356418 | OK083541 | ON052760 | OK356469 | OM883852 |
| OK356422 | OK083539 | ON052759 | OK189656 | MZ702716 |
| OK067256 | OK190637 | ON052771 | OK190626 | MZ702744 |
| ON052762 | OK356425 | MZ558154 | ON052767 | OM883999 |
| OK077992 | OK083539 | OM884031 | OK189639 | OM846619 |
| OK085481 | MZ359842 | MZ558134 | OK356470 | OK083546 |
| OL966477 | OK083540 | MZ544375 | OK189618 | OM884059 |
| OK190652 | ON052766 | OK356453 | OM900048 | OM884005 |
| OK083544 | OK356411 | OK356458 | OL966478 | MZ702529 |
| OK356414 | ON052761 | OK356431 | OK189647 | OM900055 |
| ON052768 | OK190619 | MZ702718 | OK037152 | OM900049 |
| OM899729 | OK190635 | OK067247 | MZ402742 | OM884044 |
| OK189646 | OK190635 | OK356464 | MZ359842 | OM900055 |
| OM884064 | OK189630 | OK189645 | OK356414 | OM884065 |
| OK190620 | OK356432 | OK190627 | OK061055 | OM884048 |
| OK085263 | OK189631 | ON052768 | OP781960 | OM884004 |
| OL966478 | OM884012 | OK356421 | OK190632 | OK356420 |
| OK189646 | MZ702638 | OK061055 | OK189642 | OM883928 |
| OK356455 | OK356468 | MZ544375 | OK189635 | OM883999 |
| OK189619 | OK356424 | OK189651 | OK190647 | MZ702716 |
| OK356426 | MZ702482 | MZ558096 | OL456172 | OK190646 |
| OK189616 | OK190613 | OK189623 | OM884057 | OK356457 |
| MZ713389 | OK085466 | OK356433 | OK356467 | OK356457 |
| OK356423 | OK356419 | ON052759 | ON052758 | OM884061 |

Omicron Variant (N=153):

| OQ852499 | OQ852564 | OQ852550 | ON063244 | OP295702 |
| --- | --- | --- | --- | --- |
| OQ852542 | OQ852568 | OP295722 | OQ852591 | OK189612 |
| OQ852574 | OQ852602 | OQ852571 | OP295703 | OP295749 |
| OQ852537 | ON052756 | OQ852576 | OQ852586 | OV376348 |
| OM125952 | OQ852507 | OP295704 | OQ852526 | OP295731 |
| OQ569692 | ON063250 | OQ852596 | OQ852552 | OQ852500 |
| OM236440 | OQ852549 | OP295712 | OQ852594 | OP295735 |
| OP295714 | OQ569708 | OM226178 | OM090482 | OQ569690 |
| OP295725 | OQ569698 | OQ852546 | OQ852533 | OP295718 |
| ON052754 | OQ852532 | OQ852527 | OQ852555 | OQ569694 |
| OQ852562 | OP295720 | OQ852584 | OP295726 | OP295757 |
| OQ852536 | OQ852530 | ON063248 | OQ852572 | OQ852538 |
| OK189610 | OQ852545 | OQ852603 | OQ852608 | OV330961 |
| OQ852510 | OQ852521 | OP295747 | OQ852496 | OQ852540 |
| OQ852566 | OM264106 | OV389241 | ON052769 | OM156281 |
| OQ852516 | OK189621 | OQ852599 | OQ852534 | OQ852606 |
| OQ852539 | OQ852517 | OQ852508 | OM099891 | ON052775 |
| OQ852504 | OQ852553 | OM198122 | OM085839 | OQ852551 |
| OQ569704 | OQ569688 | ON063248 | OP295730 | OM050947 |
| OP295745 | OQ852560 | OQ852506 | OQ569689 | OQ852563 |
| OQ569707 | OP295738 | OQ852503 | ON052764 | OQ852547 |
| OQ852598 | OM261479 | OP295721 | OP295711 | OQ852578 |
| OQ569710 | OQ852581 | OQ852558 | OP295729 | OQ569693 |
| OQ852559 | OQ852554 | OM197805 | OQ852548 | OM199444 |
| ON052765 | OQ852590 | OP295728 | OQ852523 | OQ852509 |
| OQ852543 | OQ852514 | OQ852525 | ON063242 | OQ852511 |
| OP295709 | ON063251 | OQ569712 | OM119018 | OQ852604 |
| OM125932 | OQ852529 | ON052753 | OQ852524 | OK189650 |
| OM229143 | OQ852498 | OQ852573 | OQ852512 | OQ852561 |
| OQ852505 | OQ852582 | ON052755 | OP295705 |  |
| OQ852607 | OQ852535 | OQ852593 | OQ852501 |  |
